# MiRNA encoded PTEN’s Impact on Clinical-Pathological Features and Prognosis in Osteosarcoma: a Systematic Review and Meta-Analysis

**DOI:** 10.1101/2024.05.16.24307477

**Authors:** Edward Kurnia Setiawan Limijadi, Robin Novriansyah, Danendra Rakha Putra Respati, Kevin Christian Tjandra

## Abstract

**Background:** Osteosarcoma (OSC) is considered one of the most common malignant bone tumours in adolescents. Due to OSC’s poor prognosis, a comprehensive approach to exploring these aspects is highly needed to improve the survival probability of OSC. In this study, we tried to explore the significance of miRNA-encoded PTEN for clinical-pathological features and prognostic value in OSC.

**Method:** We performed this systematic review and meta-analysis using articles and sources published between 2013 and 2023 from six databases (Scopus, PubMed, ProQuest, Science Direct, Sage Pub, and Cochrane). Included studies were clinical cross-sectional studies. Other study designs, articles not written in English, without full text, and not relevant—were excluded. Then, ROBINS-I is used to evaluate the distance. The results are constructed according to the PICOS criteria in a table. The expression of miRNA related to OSC is assessed in the meta-analysis as the main outcome to determine its ability as a diagnostic and prognostic agent for OSC. This systematic review followed the PRISMA guidelines.

**Results:** A total of 17 studies were included in the final screening. The meta-analysis showed significantly increased (p < 0.00001) miRNA expression in patients with osteosarcoma compared to healthy controlled with pooled md (2.85) (95% CI: 2.69, 3.02; I^2^= 22%, p= 0.20), the high inverse correlation (p < 0.001) between miRNA and PTEN expression was shown as mean effect size (−0.681) (95% CI: −0.787, −0.536; I^2^ = 75%, p < 0.0001), and the prognostic evaluation of overall survival was significantly increased in low expression miRNA (p < 0.00001) with pooled OR.

**Conclusion:** Fiveteen miRNAs from 17 studies were found, and together with PTEN expression, they may serve as potential prognostic biomarkers for OSC. High-level levels of miRNA expression are correlated with low PTEN expression, leading to a bad prognosis for OSC.

## 1. Introduction

Osteosarcoma (OSC) is the most frequent bone malignancy in both children and adults. It is a malignant mesenchymal tumour that accounts for 20–40% of all occurrences of bone malignancies. The metaphysis of the long bones in the lower extremities is the most frequent site of OSC, which is most common in people between the ages of five and twenty-five [1]. The inferior extremities, particularly the distal femur, proximal tibia, and proximal humerus, are shown to be the site of 74.5% of cases. In the meantime, factors like as age, gender, tumour location, biomarker levels, onset, and metastatic presence all affect the prognosis of OS. 61±15% is the 5-year relative survival rate for all age groups, varying based on disease onset and gender. However, this figure drastically decreases to 20±5% in patients with metastasis. Approximately 10-20% of OSC patients experience metastasis, with the lungs being a common site of involvement [2].

The high fatality rate drives the need for a more comprehensive multidisciplinary approach to the diagnosis and management of OSC. To date, several treatment options for OSC include surgical excision, radiotherapy, and multi-agent systemic therapy. Several evaluations are required to ascertain which therapy will yield the best response in the patients. Furthermore, invasive biopsy is still required for OSC diagnosis evaluation and confirmation [3,4]. However, due to variations in sample collection, the accuracy of the biopsy-derived diagnosis and prognosis can fluctuate. Therefore, the diagnostic approach is an important aspect to develop to achieve better prognosis, and one method with great potential is identifying biomarkers [5–7].

Biomarkers themselves can be used as therapeutic, prognostic, and diagnostic agents. MicroRNAs (miRNAs) are a class of endogenous non-coding RNAs with lengths of 19-25 nt. They are involved in differentiation, cell death, and the cell cycle by regulating gene expression through direct inhibition and degradation of mRNA translation [8]. In malignancies, this inhibitory function makes them tumor suppressors and biomarkers for the diagnosis and prognosis of patients. PTEN is one of the major tumor suppressors that has been extensively studied for its effects on cancer development, such as breast cancer, prostate cancer, and lung cancer. PTEN (phosphatase and tensin homolog deleted on chromosome ten) is a candidate tumor suppressor located on chromosome 10q23. PTEN works in the PI3K/AKT regulatory pathway, a lipid kinase family axis with the PIP3 product, which is involved in controlling cell proliferation to prevent cell development into cancer. The increase and decrease in PTEN expression are regulated by a number of proteins in posttranslational, posttranscriptional, and transcriptional mechanisms. One of these proteins is miRNA, which suppresses PTEN expression posttranscriptionally. In this systematic review and meta-analysis, we summarize current knowledge on the correlation between miRNA-PTEN and its impact on the clinicopathological and prognostic aspects of OSC, which has not been previously explored within the scope of a wide range of articles.

## 2. Methods

### Eligibility criteria

The Preferred Reporting Items for Systematic Reviews and Meta-Analyses (PRISMA) were used for this systematic review [9]. From 2013 to 2023 (last search date September 20^th^, 2023), we have included original content. The study included original research publications that met the inclusion criteria for autologous clinical cross-sectional. Technical reports, editor’s responses, narrative reviews, systematic reviews, meta-analyses, non-comparative studies, in silico studies, in vitro studies, in vivo studies, applied Scientific posters, research proposals, and conference abstracts were all diminished. Articles not written in English, with incomplete content, and not related to the miRNA and PTEN Gene expression in correlation to OSC as well as survival rate were also eliminated. The desired PICO criteria of the selected products included i) patients with OSC; ii) miRNA and PTEN expression evaluation towards OSC patients and its survival rate; iii) Comparison, normal cells either obtain from the same patients or other normal patients; and iv) Results, miRNA and PTEN evaluation utilizing RT-qPCR and two years survival rate.

### Data search and selection

Research for this study were gathered through Scopus, PubMed, ProQuest, Science Direct, Sage Pub, Cochrane database searches. We used the following combined keywords to capture all potentially eligible literature in each databases: “(osteosarcoma) AND (miRNA OR microRNA) AND (PTEN) AND (clinicopathology OR prognosis)”. The database was searched from its establishment until September 20^th^ 2023 during 10-year periods prior to this review. The boolean operator was utilized among the Medical Subject Headings (MeSH) keywords determined from national institute of health (NIH) national library of medicine browser. The studies were kept using Mendeley Group Reference Manager in the authors’ library. Clinical type articles were used as a filter in the Pubmed database, the the research article type was used for the Scopus, PubMed, ProQuest, Science Direct, Sage Pub, Cochrane database filter. All of these processes for article selection were conducted by the two independent authors (DRPR, KCT) who performed the literature searching process. The identified articles from those databases were first filtered according to their titles and abstracts, and then duplicates were eliminated. The same two authors conducted a second round of full-text evaluations for these publications that made it past the first screening round to determine whether they met our inclusion/exclusion criteria. All differences were addressed by engaging in discussions with the other author (EKSL, RN), who assessed the suitability of studies for inclusion in the synthesis.

### Data extraction

After the final screening, the pertinent information from studies was retrieved and entered into a Google Spreadsheet. Recorded data in characteristic table consited of author, year, country, study design, sample size, mean age, miRNA type, miRNA expression with poor prognosis (high or low), and cut-off. All of the data extraction was done by two independent author (DRPR, KCT). The primary outcome of this study consisting of the survival analysis, miRNA/PTEN correlation, and clinicopathology features (TNM stage, metastasis, and gender). The desired outcome for the survival analysis was 5-year Overall Survival (OS), defined as the time from the initiation of therapy until death from any cause. Using hazard ratios (HR), the impact of miRNA expressions on survival was evaluated. If given by the authors, a univariate HR estimate and 95% confidence intervals were taken straight from each study. If not, the suggested approach was used to extract the p values of the log-rank tests, 95% confidence intervals, the number of events, and the number of patients at risk in order to estimate the HR [10]. In cases where the number of events and patients at risk was not reported, we use Engauge Digitizer version 4.1 to reconstruct the kaplan-meier curves, then we calculated the HR using the same method as before. In patients with aberrant miRNA, a pooled HR < 1 suggested a better prognosis, whereas a pooled HR > 1 suggested a worse prognosis.

### Study Risk of Bias Assessment (Qualitative Synthesis)

Risk-of-bias tool used to assess the bias of included studies was the ROBINS-I from Cochrane Collaborations, was used by three independent author (KCT, DRPR, RN). Any disagreements regarding the bias assessment were discussed further and settled between the 3 reviewers. The result is shown in **Figure 2**. The authors’ evaluations were categorized as "low risk," "high risk," or "some concerns" of bias. For analyzing the quality of included studies, we used Newcastle-Ottawa Scale (NOS) from the Ottawa Hospital Research Institute (OHRI), a quality appraisal tool for cohort and crossectional studies which includes 3 domains of assessment: (1) selection of participants; (2) comparability between exposed and non-exposed cohort; and (3) outcome ascertainment [11]. For cross-sectional research, the maximum possible score ranges from 0 to 10, while for cohort studies, it ranges from 0 to 9. Articles with a score of at least 7 were deemed to have "good" quality. To maintain the validity of the data included in the current investigation, papers that were judged to have a high risk of bias will be eliminated from the systematic review.

### Statistical Analysis

The results were displayed in the characteristic table as, mean difference (MD) with standard deviation (SD) for miRNA expression with poor prognosis, HR with a 95% confidence interval (CI) for OS, and correlation coefficient (r) with a 95%CI for miRNA/PTEN expression. Gender, TNM stage, and metastatic status were presented as odd ratios (OR) with 95% CI. For pooled analysis, we converted the data expressed as median and interquartile range (IQR) or as median, minimum, and maximum into mean and standard deviations (SD) using a combination formula from Luo D et al. and Wan X et al [12,13]. The I-squared (I2) statistic was employed in this review to evaluate the heterogeneity amongst studies, with I2 values greater than 50% classified as significant heterogeneity.When the included studies were deemed homogeneous (little variability in study results or variation owing to random error), as shown by an *P* ≥ 0.10 or *I*2 ≤ 50%, meta-analysis was performed using a fixed-effect model (FEM). Otherwise, we used a random-effect model (REM) if *P* < 0.10 or *I*^2^ > 50%. The pooled mean difference estimate was presented in a forest plot. When there were more than 10 studies on each outcome of interest, a publication bias analysis was conducted using the funnel plot and Egger’s test; if an asymmetry in the funnel plot was discovered, we planned to review the characteristics to determine whether the observed asymmetry could be attributed to publication bias or alternative factors such as methodological heterogeneity among the studies. All of the analyses were carried out using Comprehensive Meta-Analysis software, which can be accessed at https://meta-analysis.com/.

## 3. Result

### Study Selection

A total of 12,939 studies were found in the database (5622 from Scopus, 31 from Pubmed, 5311 from Proquest, 1720 from Science Direct, 116 from Sage Pub, and 156 from Cochrane). Then, 4842 studies were imported to the Mendeley Group Reference Manager in the authors’ library after the aforementioned criteria were added, and this was done before the selection procedure. The number of 4312 studies was eliminated from this process due to irrelevant topics. Eventually, 561 studies were screened manually by the authors. Out of all, 530 studies, they did not provide sufficient data to answer the research question, and because their study designs did not meet the criteria for inclusion (clinical cross-sectional), two studies were removed due to duplication, and three were removed due to irretrievable full text. The retrieval of the complete text for the remaining twenty-six studies was then tested. After the full text reading was done, only seventeen articles were considered eligible to be obtained as a result. The flow chart for the PRISMA diagram illustrating our research selection procedure is shown in **Figure 1**. Using the Cochrane ROBINS-I tool, the seventeen included studies were evaluated for eligibility. All seventeen of the listed studies from this process passed the evaluation bias check. The PRISMA flow chart contained records of the research selection procedures.

**Figure 1.**
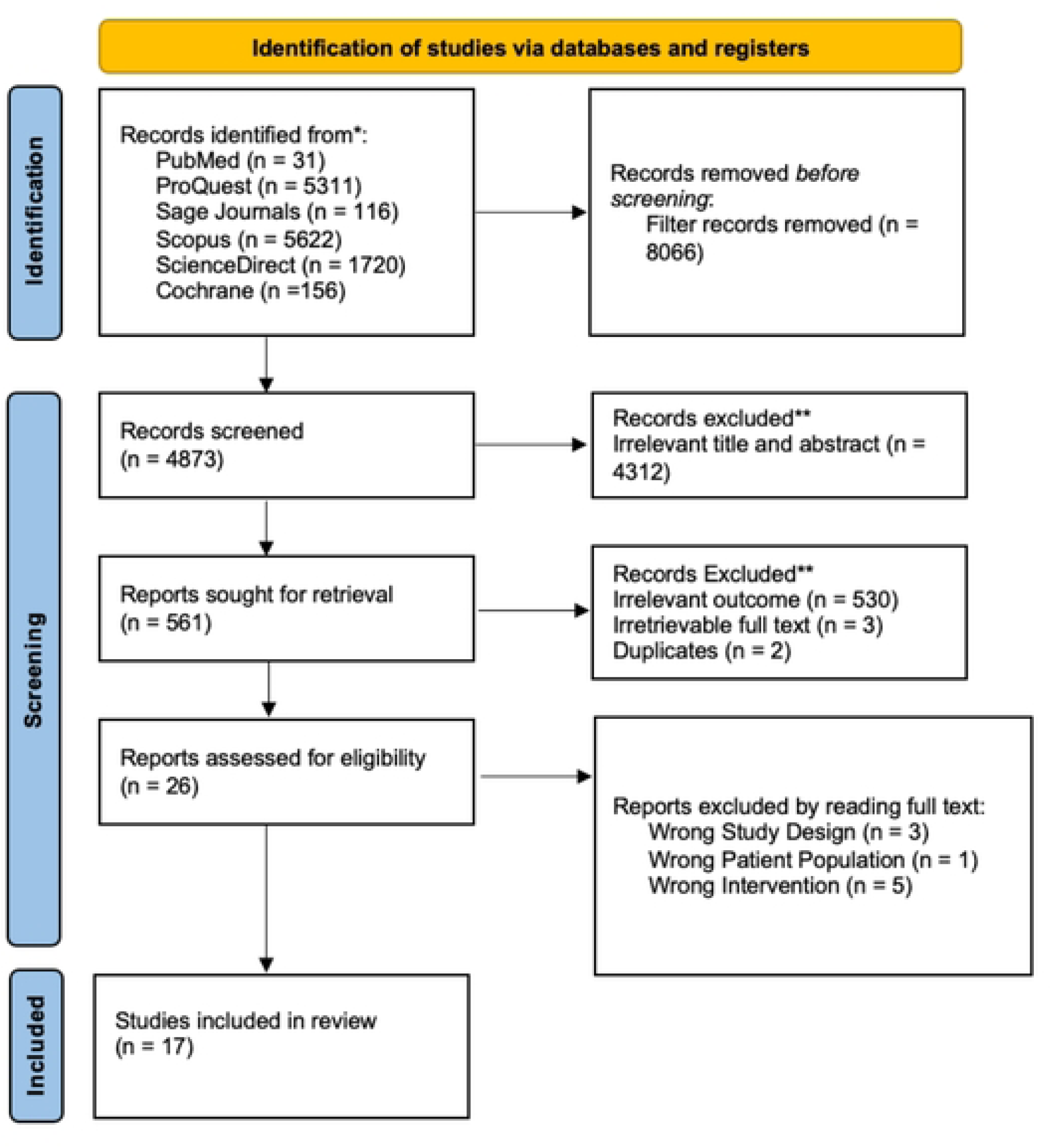
PRISMA 2020 flow diagram.

### Study Characteristics

The seventeen studies that made up this systematic review comprised a total of 760 participants. These each four studies were conducted in different countries. Studies were carried out in Japan, South Korea, France, China, USA, Iran, Singapore, and Spain. The complete study characteristics, including the PICO of each study, are stated in **Table 1**.

**Table 1.** Study Characteristic.

| No | Author | miR type | Sample type | Sample size |  | Cut off | MiRNA expression | Assay method |
| --- | --- | --- | --- | --- | --- | --- | --- | --- |
|  |  |  |  | miRNA high | miRNA low |  |  |  |
| 1 | Zhao D., et.al.;2017 | miRNA-19a | Tissue | 25 | 25 | NR | upregulated | RTq-PCR |
| 2 | Chen J., et.al.;2015 | miRNA-130a | Tissue | 86 | 86 | median | upregulated | RTq-PCR |
| 3 | Yuan H., et.al.;2015 | miRNA-1908 | Tissue | 46 | 9 | NR | upregulated | RTq-PCR |
| 4 | Hu X., et.al.;2018 | miRNA-21 | Tissue | 46 | 20 | NR | upregulated | RTq-PCR |
| 5 | Liu Q., et.al.;2018 | miRNA-29 | Tissue | 60 | 60 | median | upregulated | RTq-PCR |
| 6 | Liu J., et.al.;2015 | miRNA-214 | Tissue | 22 | 22 | NR | upregulated | RTq-PCR |
| 7 | Gao Y., et.al.;2014 | miRNA-17 | Tissue | 28 | 28 | NR | upregulated | RTq-PCR |
| 8 | Zhuang M., et.al.;2018 | miRNA-524 | Tissue | 20 | 20 | NR | upregulated | RTq-PCR |
| 9 | Sun C.,et.al.;2022 | miRNA-181a-5p | Tissue | N/A | N/A | NR | upregulated | RTq-PCR |
| 10 | Zhang H.,et.al.;2016 | miRNA-148a | Tissue | 92 | 92 | median | upregulated | RTq-PCR |
| 11 | Zhu J.,et.al.;2015 | miRNA-221 | Tissue | 16 | 12 | NR | upregulated | RTq-PCR |
| 12 | Yu W.,et.al.;2020 | miRNA-744 | Tissue | 25 | 25 | NR | upregulated | RTq-PCR |
| 13 | Fu Y.,et.al.;2020 | miRNA-208a-3p | Tissue | 10 | 10 | NR | upregulated | RTq-PCR |
| 14 | Xiao J.,et.al.;2017 | miRNA-92a | Tissue | 68 | 68 | median | upregulated | RTq-PCR |
| 15 | Yu L.,et.al.;2017 | miRNA-214 | Tissue | 15 | 15 | NR | upregulated | RTq-PCR |
| 16 | Tian Z.,et.al.;2014 | miRNA-128 | Tissue | 100 | 100 | median | upregulated | RTq-PCR |
| 17 | Zhao H., et.al.;2019 | miRNA-21<br>miRNA-221 | Tissue | NR<br>NR | NR<br>NR | median | upregulated | RTq-PCR |

### Risk of Bias in Studies

Each cohort and cross-sectional study underwent a thorough assessment of its quality using the ROBINS-I risk-of-bias method. In nine of the investigations, there were four studies identified as a study with some concern of bias due to unclear randomization process between the allocation of intervention and control groups. An overview of the bias risk assessment is shown in **Figure 2**.

**Figure 2.**
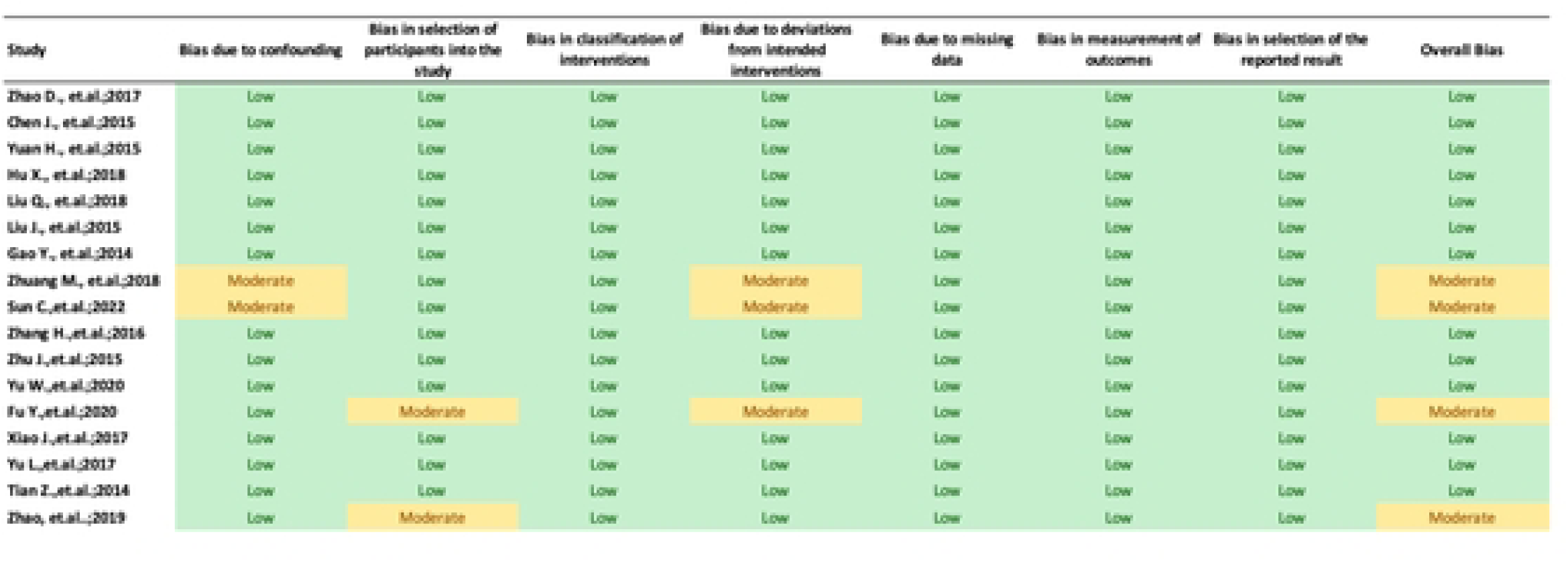
Risk of Bias Assessment

### Clinicopathologies

A meta-analysis of five studies was used to evaluate the gender clinicopathologic relationship between OSC overexpression and miRNA encoding PTEN. A non-significant effect (P = 0.28) with a pooled odds ratio (OR) of (1.24) (95% CI: 0.84, 1.83) is seen in **Figure 3** forest plot. This outcome suggests that events do not differ for males and females. Additionally, the forest plot demonstrated heterogeneity for the gender odds ratio clinicopathology (I^2^ = 0%; P = 0.53). **Figure 4** displays an analysis of the metastatic clinicopathologic associated with miRNA encoding PTEN for OSC from three journals. A significant effect (P < 0.00001) was seen in the forest plot, with a pooled OR of 0.24 (95% CI: 0.13, 0.43). The combined OR showed that an increase in the events leading to OSC metastasis was caused by over-expression of the miRNA encoding PTEN. The results showed heterogeneity (I^2^ = 12%; P = 0.32). The assessment of OSC development entailed a review of its TNM staging concerning miRNA encoding PTEN, as shown in **Figure 5**. A significant effect (P = 0.02) can be seen in the forest plot, where the pooled odds ratio (OR) was 0.59 (95% CI: 0.38, 0.93). The cumulative OR indicated that an unfavourable prognosis for OSC was linked to over-expression of miRNA encoding PTEN. Heterogeneity was notably found (I^2^ = 11%; P = 0.35).

**Figure 3.**
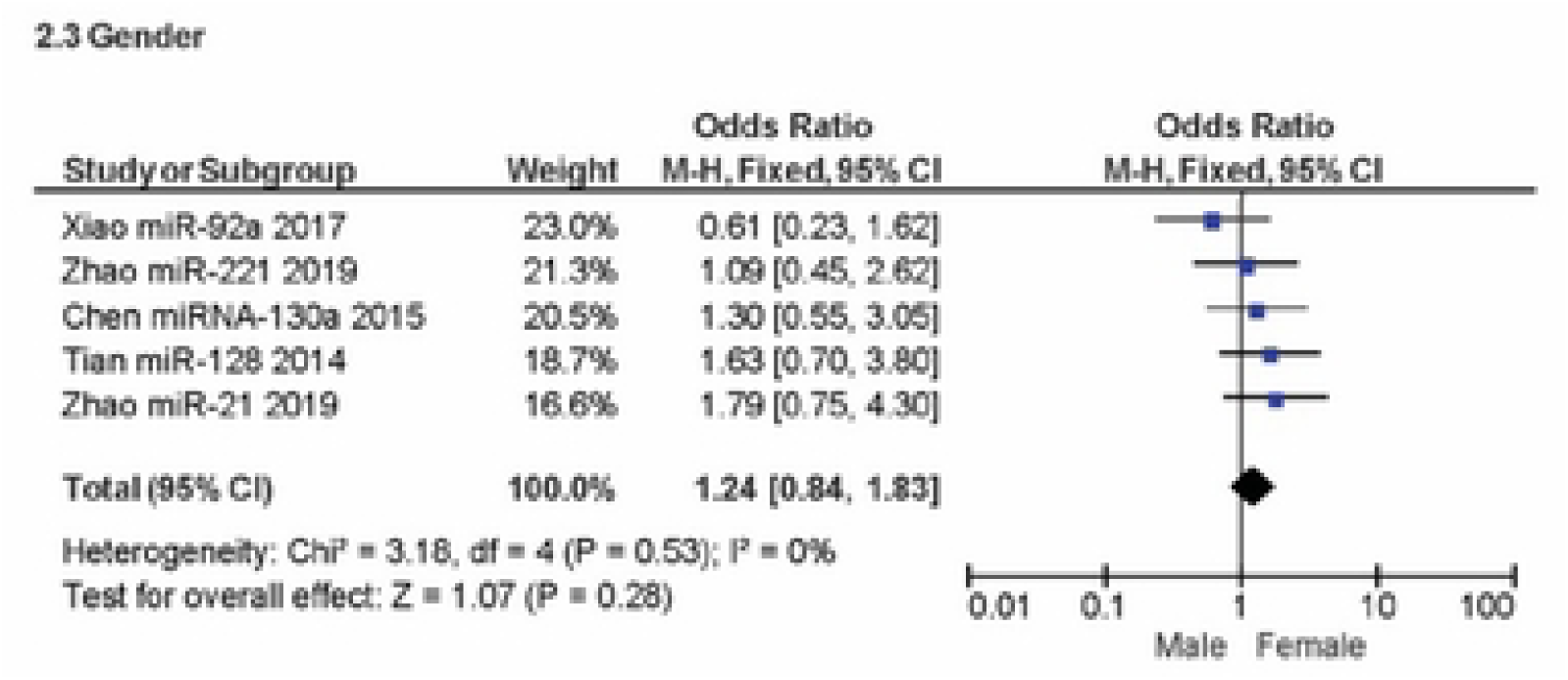
Forest plot of gender-related to miRNA overexpression

**Figure 4.**
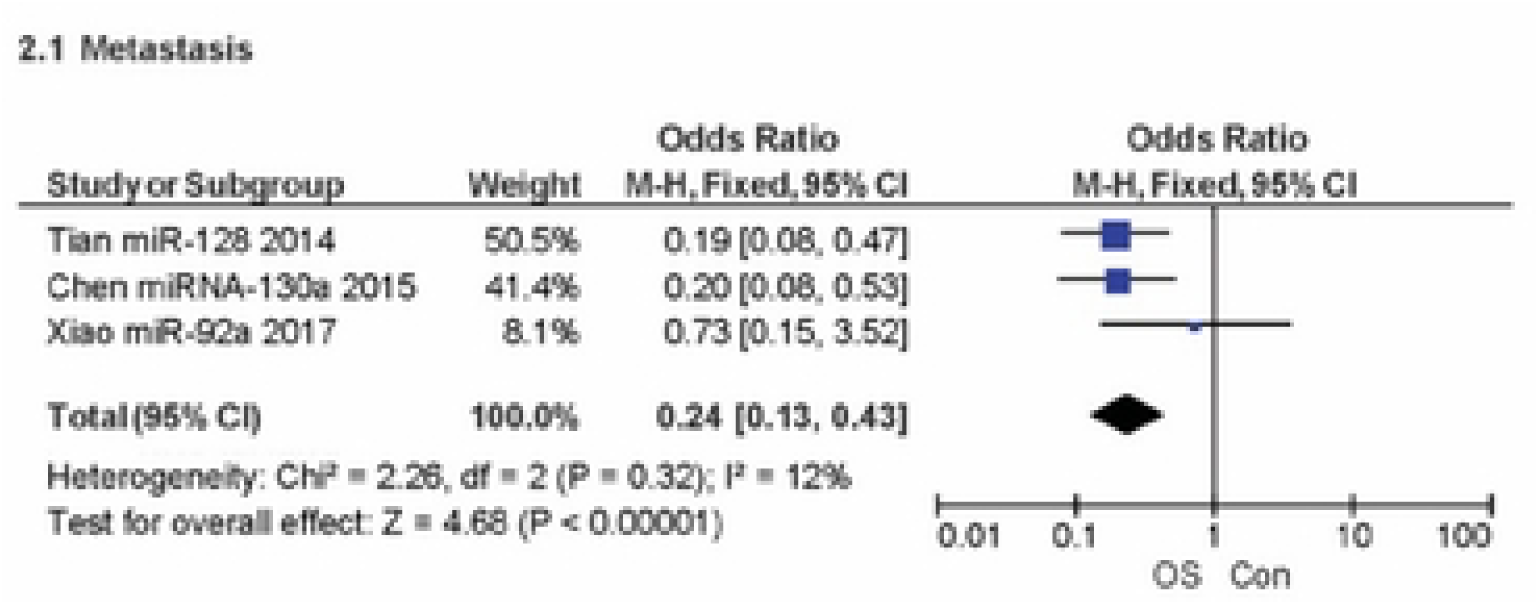
Forest plot of metastasis-related to miRNA overexpression

**Figure 5.**
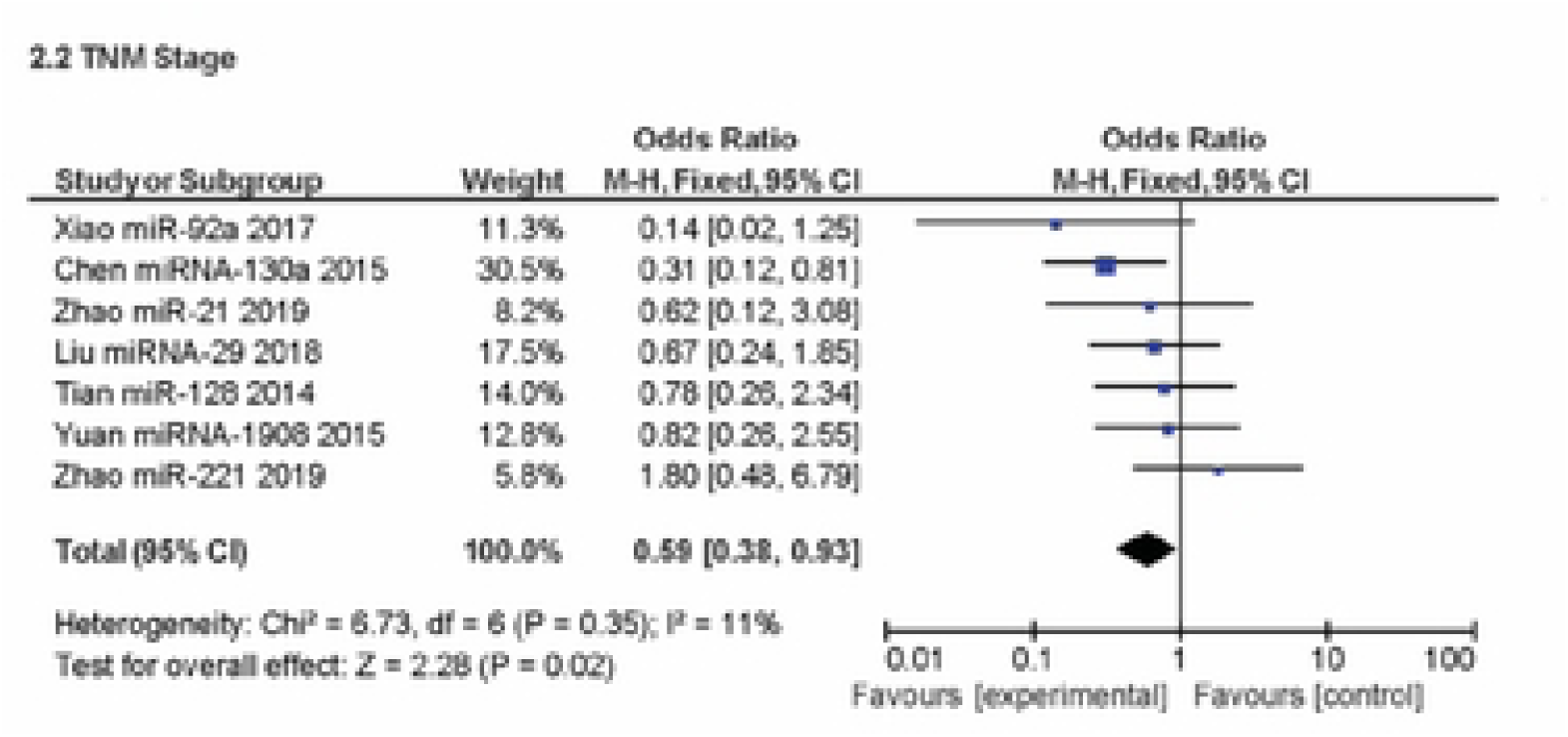
Forest plot of TNM staging related to miRNA encoding PTEN overexpression

### Prognostic survival analysis

The correlation between OS rate and miRNA overexpression was thoroughly examined in this study. The assessment, depicted in **Figure 6** through meta-analysis, revealed a substantial effect (P < 0.00001), with a pooled Hazard Ratio (HR) of −12.38 (95% CI: −13.75, −11.01). The aggregated HR underscored that OSC patients exhibiting miRNA encoding PTEN overexpression face a mortality risk more than twice as high. Noteworthy heterogeneity was observed (I^2^ = 20%; P = 0.27).

**Figure 6.**
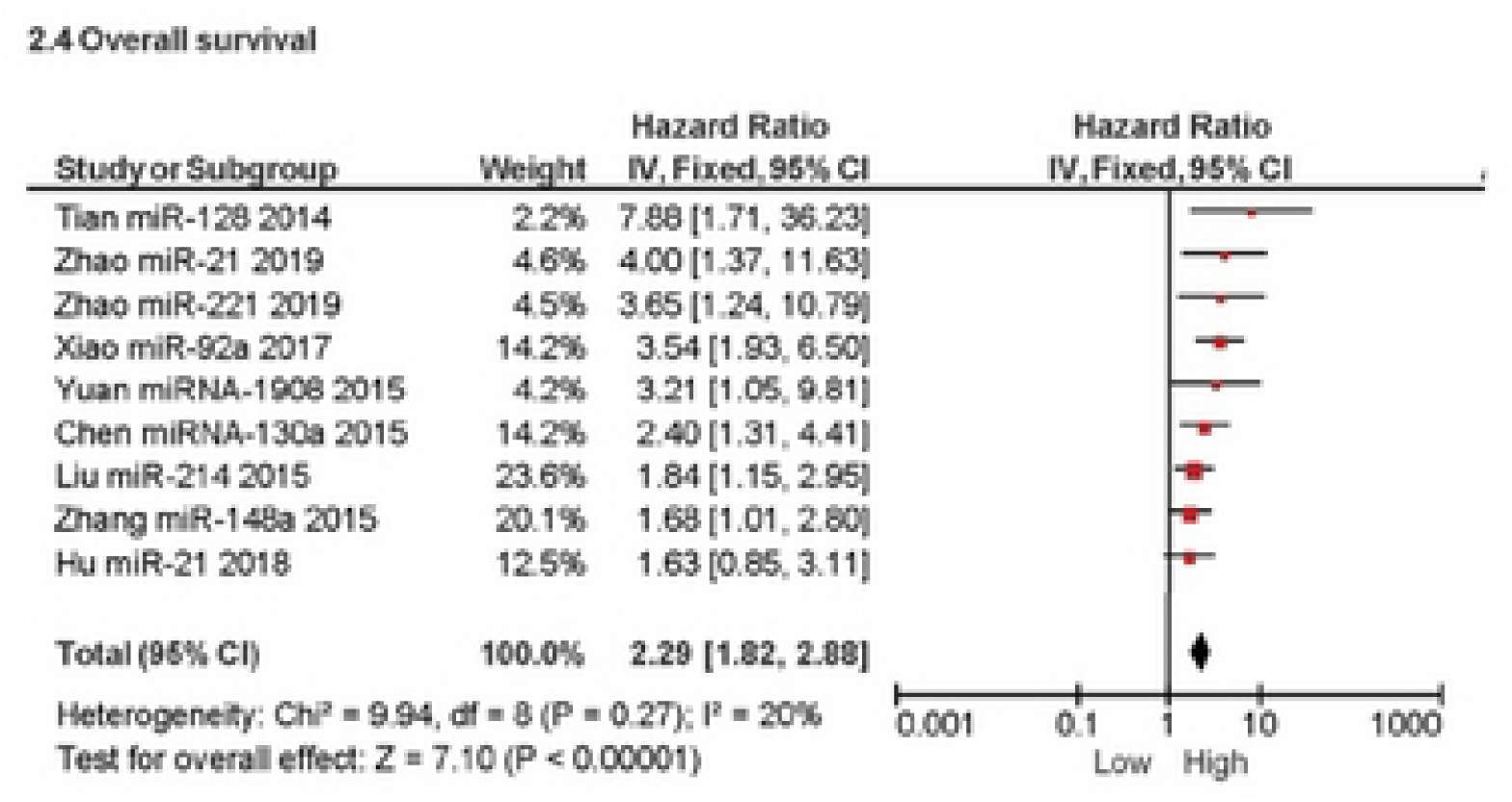
Forest plot of overall survival related to miRNA encoding PTEN overexpression

### miRNA and PTEN correlation for osteosarcoma expression

A meta-analysis comparing the population with OSC to the healthy population was conducted as part of the inquiry into miRNA overexpression. A significant effect (P < 0.00001) was seen in the forest plot, as shown in **Figure 7**, with a pooled mean difference (MD) of 2.83 (95% CI: 2.65, 3.02). When comparing the expression of miRNA in patients with OSC to that in healthy individuals, the combined MD showed a substantial increase in expression. Heterogeneity was found, notably (I^2^ = 27%; P = 0.16). Meanwhile, this meta-analysis examined PTEN expression and its relationship to miRNA expression. This result was shown in **Figure 8**. A significant effect (P < 0.00001) was seen in the forest, with a pooled mean difference (MD) of −1.51 (95% CI: −1.68, −1.34). PTEN expression was lower in patients with miRNA overexpression, according to the pooled MD. Heterogeneity was found (I^2^ = 77%; P < 0.00001).

**Figure 7.**
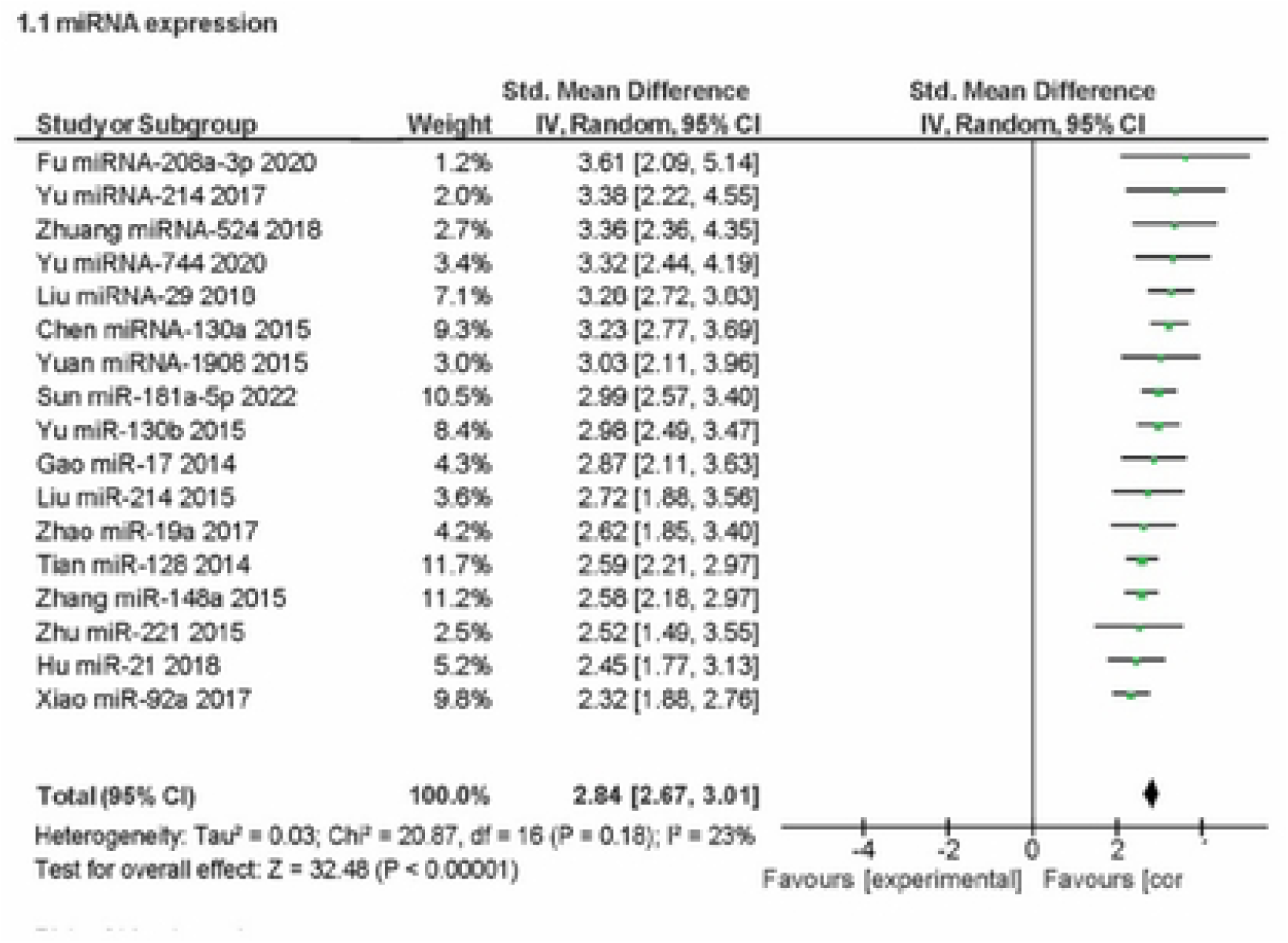
Forrest plot of high/low miRNA expression

**Figure 8.**
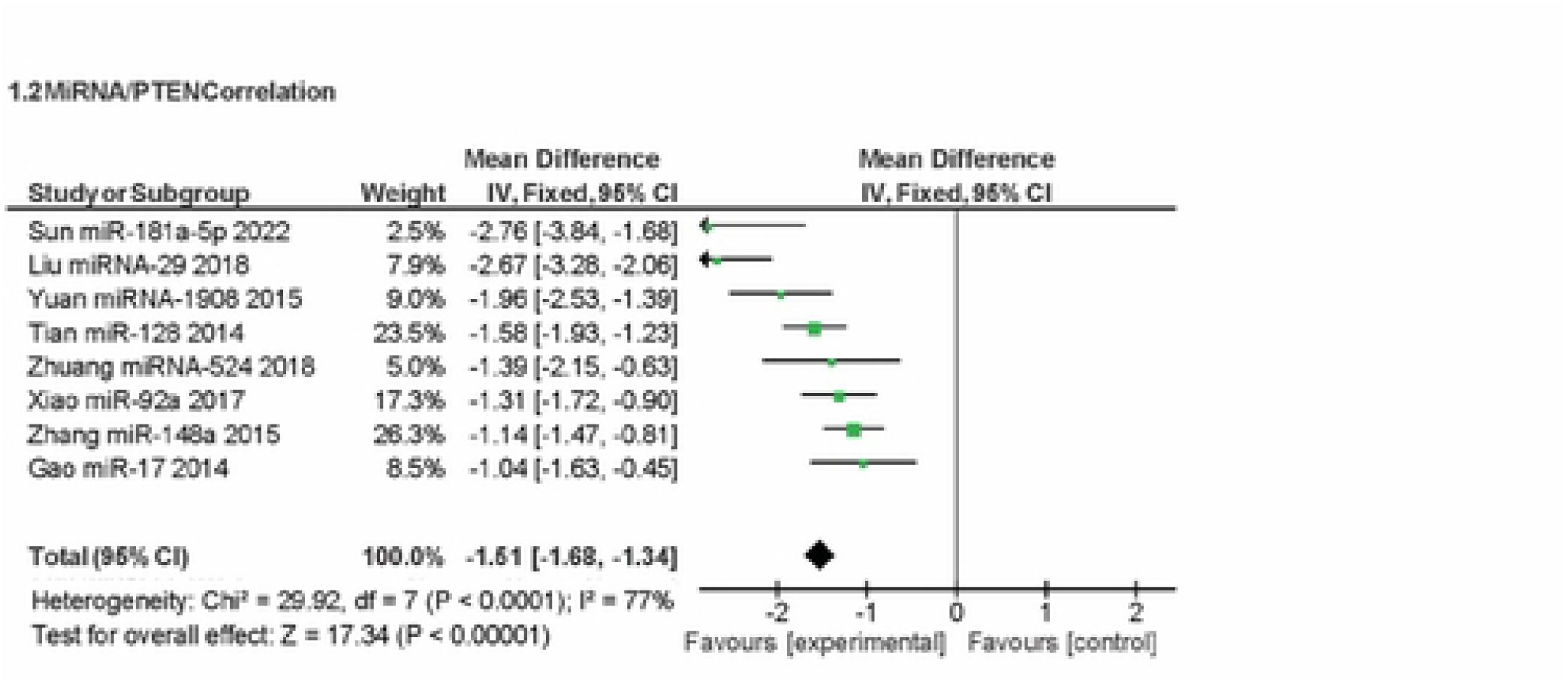
Forrest plot of miRNA and PTEN expression correlation

### Reporting biases

Egger’s test and Begg’s funnel plot were used to assess the meta-analysis’s publication bias. The funnel plot among the 13 research did not clearly demonstrate any signs of asymmetry, as **Figure 9** illustrates. Furthermore, Egger’s test in the meta-analysis indicated no evidence of publication bias (P>0.05).

**Figure 9.**
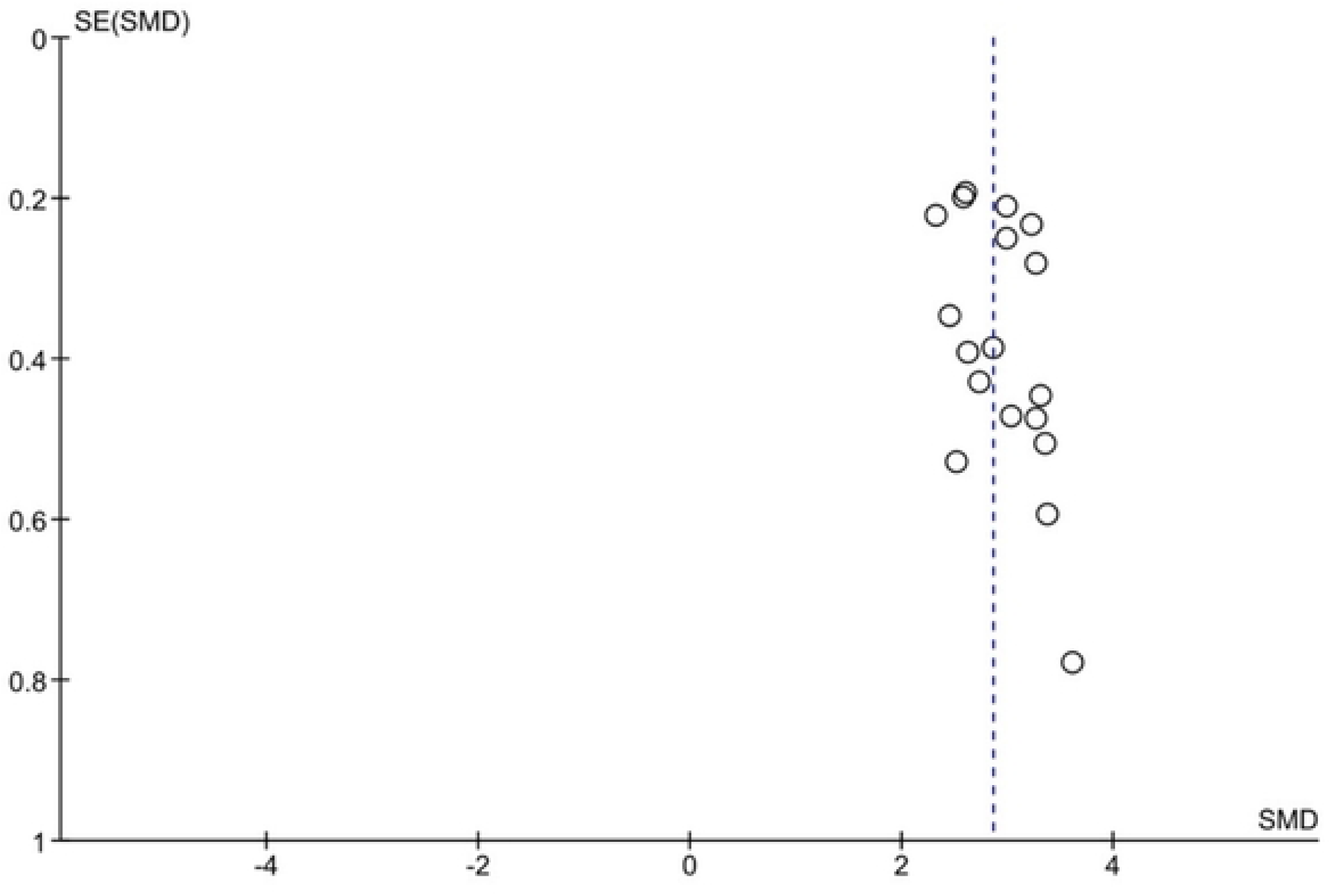
Funnel plot of high/low miRNA expression

## 4. Discussion

The identification of Phosphatase and tensin homolog deleted on chromosome ten (PTEN) dates back to 1997. It marked the inaugural recognition of a tumor suppressor gene possessing tyrosine phosphatase activity. The nomenclature "phosphatase and tensin homolog deleted on chromosome ten" was derived from its location at 10q23 [14]. Crucial to the principal regulatory pathway of cell growth is phosphatidylinositol 3,4,5-trisphosphate (PIP3), capable of stimulating cell growth and initiating tissue cell apoptosis [15]. PTEN intervenes by dephosphorylating one of the three phospho-groups of PIP3, modulating the cell growth pathway and prompting cellular self-destruction, thereby instigating abnormal cell death[16]. Furthermore, PTEN’s tumor suppressor role extends to cell cycle regulation, where it fosters p27Kip1 binding to the CyclinE/cyclin-dependent kinase 2 (CDK2) complex, inhibiting CDK2 kinase activity. This inhibition prevents cells from entering the S phase and correlates with the down-regulation of RB protein phosphorylation levels [17].

PTEN down-regulation has been documented in various malignant tissues such as glioma, endometrial cancer, lung cancer, and prostate cancer [8,18,19]. Previous investigations into the impact of PTEN expression on OSC patient prognosis yielded conflicting results. For instance, Sun et al. [16] posited that positive PTEN expression is unrelated to gender, age, tumor size, and metastasis, while Han et al. and Xie et al [20–22]. reported an association between PTEN expression and OSC metastasis. In this comprehensive report, a meta-analysis encompassing all available studies on PTEN expression and OSC patients was conducted to elucidate its relationship with the prognosis of OSC.

This study demonstrates that PTEN is a direct target of multiple miRNAs, including miRNA-19a, miRNA-130a, miRNA-1908, miRNA-21, miRNA-29, miRNA-17, miRNA-524, miRNA-181a-5p, miRNA-148a, miRNA-221, miRNA-744, miRNA-208a-3p, miRNA-92a, miRNA-214, and miRNA-128, which are overexpressed in OSC tissues compared to normal tissues. PTEN shows a negative correlation with miRNA expression in OSC tissue. Studies by Gao Y. (2014) demonstrated PTEN as a target of miRNA-17, suppressing the WT 3’-UTR in HEK293 cells [23]. Yuan H (2015) reported that miR-1908 overexpression reduces PTEN expression, confirmed through luciferase activity comparison in OSC cells transfected with miRNA-1908 [24]. Xiao J. (2017) reported increased miR-92a expression reduces PTEN levels, leading to increased expression of p-AKT(Ser473), mTOR, p-p27(Thr157), and p-MDM2(Ser166) in MG-63 cells, indicating miR-92a regulates the PTEN/AKT pathway in OSC cells [25].

Previous studies have detailed the post-transcriptional regulation of oncogenes and tumor suppressors by microRNAs (miRNAs), involving epigenetic mechanisms such as DNA methylation, chromatin modification, and non-coding RNAs (ncRNAs), including miRNAs and long non-coding RNAs (lncRNAs). MiRNAs targeting PTEN primarily focus on the slender region of the 3’ untranslated region (3’UTR), leading to the downregulation of PTEN expression [8,19]. Consequently, the overexpression of miRNAs suppresses PTEN function in the PI3K/Akt pathway, promoting OSC growth [26,27]. In tumor cells, PTEN acts as an anti-proliferative agent by inhibiting cyclin D1 transcription through AKT inactivation and increasing lipid phosphatase activity in the cytoplasm, resulting in elevated p27 expression. PTEN also mediates apoptosis through the activation of caspase-3 and TP53 [28]. Furthermore, PTEN regulates the Epithelial Mesenchymal Transition (EMT), an early stage in the metastasis cascade [29,30]. Thus, PTEN indirectly influences the prognosis of OSC and other tumors, such as breast, kidney, and lung cancers [31–33].

We further analyzed the correlation between the expression of these miRNAs and the prognosis and clinicopathological features of OSC. A comprehensive systematic review and meta-analysis clarified the prognostic value of miRNAs and PTEN in OSC. An increase in miRNAs targeting PTEN in OSC tissues closely correlates with worse OS Xiao J. (2017) reported that OS tissues with overexpression of miRNA-92a have worse overall and event-free survival (EFS) [25]. Detailed confirmation of the role of miRNAs in OSC prognosis was provided by Zhao H. (2019), comparing miRNA-128-high/PTEN-low, miRNA-128-low/PTEN-high, and miR-128-low/PTEN-low groups, showing that upregulation of miRNA-128 and downregulation of PTEN constitute the group with the worst prognosis and clinicopathological features [34]. Zhang H. (2016) explained their findings regarding the correlation between overexpression of miRNA-148a in OSC tissues and worse OS and clinicopathological features [35].

It is essential to note that OSC prognosis can be influenced by various risk factors beyond miRNA expression, with patient clinicopathological features also playing a role. Therefore, we evaluated the relationship between miRNA expression and OSC clinicopathological features. The relevance of each clinicopathological feature and the overexpression of miRNAs to OSC prognosis was explained in a study by Zhao H. (2019) [34]. MiRNA-21, miRNA-221, metastasis, and tumor staging were identified as major independent risk factors impacting OSC prognosis compared to other parameters. It was also indicated that the overexpression of miRNA-21 and miRNA-221 has a more significant impact on worse prognosis than metastasis and tumor staging. To further confirm these findings, we investigated the correlation of miRNAs with gender, metastasis, and OSC staging.

We found that positive miRNA expression significantly associated with female gender, metastasis, TNM staging, and poor prognosis. Chen J. (2015) suggested that overexpression of miRNA-130a promotes OSC metastasis and EMT through PTEN inhibition, confirmed by transwell assay results showing increased migration and invasion in HOS58 cells [36]. Hu X. (2018) and Zhu J. (2015) indicated that positive expression of miRNA-21 and miRNA-221 increases proliferation, invasion, and migration through PTEN downregulation, subsequently promoting metastasis [37,38].

The study exhibits several strengths, including a comprehensive approach through systematic review and meta-analysis, involving 17 studies published between 2013 and 2023 from diverse databases. Methodological rigor is maintained with the use of the ROBINS-I tool for risk assessment, and adherence to PRISMA guidelines ensures transparency. Clinically relevant outcomes are explored, shedding light on the significance of miRNAs encoded PTEN in OSC. However, limitations include a restriction to clinical cross-sectional studies, potentially limiting the diversity of evidence. Exclusion of non-English articles introduces language bias, and the temporal limitation to 2013-2023 might overlook newer developments. Heterogeneity in meta-analysis and the focus on a limited set of outcome measures may impact the generalizability and comprehensiveness of the findings. Overall, the study suggests 15 miRNAs, in conjunction with PTEN expression, as potential prognostic biomarkers for OSC, though the findings should be interpreted considering these limitations.

However, this study has several unavoidable limitations. First, the analysis is based on a set of publications. Second, some Hazard Ratios (HR) and 95% Confidence Intervals (CI) were obtained through survival curve extraction, potentially reducing study accuracy. Third, the majority of studies were conducted in the Asian region, which may affect the generalizability of the results. Fourth, some included studies had small sample sizes, potentially increasing sample bias and randomization errors.

## 5. Conclusion

Using a meta-analysis technique, we examined the clinicalpathology characteristics and prognostic usefulness of miRNA-encoded PTEN in patients with OSC in this study. In conclusion, the overexpression of miRNAs encoded PTEN contributes to the unfavourable clinicopathological features and the prognosis of OSC. These factors are primarily associated with female gender, metastasis, and advanced TNM staging. Furthermore, miRNA expression has a greater impact on the decreased OS in OSC.

## STATEMENTS AND DECLARATIONS

### Registration

On August 8, 2023, this systematic review and meta-analysis was registered to the Open Science Framework (OSF). The registration was identified as MiRNA encoded PTEN’s Impact on Clinical-Pathological Features and Prognosis in Osteosarcoma: a Systematic Review and Meta-Analysis. https://doi.org/10.17605/OSF.IO/647WF.

### Availability of data and materials

#### Underlying data

All data underlying the results are available as part of the article and no additional source data are required.

#### Reporting guidelines

**Data are available under the terms of the Creative Commons Attribution 4.0 International license (CC-BY 4.0).**

## Data Availability

All relevant data are within the manuscript and its Supporting Information files.

## Acknowledgment

None

## Ethical Approval

Not applicable

## Patient’s Consent for Publication

Not applicable

